# Comprehensive Autoantibody Profiles in Systemic Sclerosis: Clinical Cluster Analysis

**DOI:** 10.1101/2022.09.29.22280527

**Authors:** Jakob Höppner, Christoph Tabeling, Vincent Casteleyn, Claudia Kedor, Wolfram Windisch, Gerd Rüdiger Burmester, Dörte Huscher, Elise Siegert

**Affiliations:** Department of Rheumatology and Clinical Immunology, Charité - Universitätsmedizin Berlin, Berlin, Germany; Department of Pulmonology, Cologne Merheim Hospital, Kliniken der Stadt Köln gGmbH, Witten/Herdecke University, Cologne, Germany; Division of Pulmonary Inflammation, Charité - Universitätsmedizin Berlin, Corporate Member of Freie Universität Berlin and Humboldt-Universität zu Berlin, Berlin, Germany; Department of Infectious Diseases and Respiratory Medicine, Charité - Universitätsmedizin Berlin, Corporate Member of Freie Universität Berlin and Humboldt-Universität zu Berlin, Berlin, Germany; Berlin Institute of Health at Charité - Universitätsmedizin Berlin, Berlin, Germany; Institute of Biometry and Clinical Epidemiology, Charité - Universitätsmedizin Berlin, Berlin, Germany

**Keywords:** Systemic Sclerosis, Scleroderma, Autoantibodies, Primary Biliary Cholangitis, Cluster Analysis, Epidemiology

## Abstract

**Background:** Systemic sclerosis (SSc) belongs to the group of connective tissue diseases and is associated with the occurrence of disease-specific autoantibodies. Although it is still controversial whether these antibodies contribute to pathogenesis, there are new insights into the development of these specific antibodies and their possible pathophysiological properties. Interestingly, they are associated with specific clinical manifestations, but for some rarer antibodies this association is not fully clarified. The aim of this study is a comprehensive analysis of the serum autoantibody status in patients with SSc followed by correlation analyses of autoantibodies with the clinical course of the disease.

**Methods:** Serum from SSc patients was analyzed using a line blot (EUROLINE, EUROIMMUN AG) for SSc-related autoantibodies. Autoantibodies to centromere, Topo-1, antimitochondrial antibodies (AMA) M2 subunit, angiotensin II type 1 receptors (AT_1_R) and endothelin-1 type-A-receptors (ET_A_R) were also determined by ELISA. We formed immunological clusters and used principal components analysis (PCA) to assign specific clinical characteristics to these clusters.

**Results:** A total of 372 SSc patients were included. 95.3% of the patients were antinuclear antibody positive and in 333 patients at least one SSc specific antibody could be detected. Four immunological clusters could be found by PCA. Centromere, Topo-1 and RP3 all formed own clusters, which are associated with distinct clinical phenotypes. We found that patients with an inverted phenotype, such as limited cutaneous SSc patients within the Topo-1 cluster show an increased risk for ILD compared to ACA positive patients. Anti-AT_1_R and anti-ET_A_R autoantibodies were measured in 176 SSc patients; no association with SSc disease manifestation was found. SSc patients with AMA-M2 antibodies showed an increased risk of cardiovascular events.

**Conclusion:** In our in large cluster analysis, which included an extended autoantibody profile, we were able to show that serologic status of SSc patients provides important clues to disease manifestation, co-morbidities and complications. Line blot was a reliable technique to detect autoantibodies in SSc and detected rarer autoantibodies in 42% of our patients.

## 1. Introduction

Systemic Sclerosis (SSc), also called scleroderma, is a rare autoimmune-mediated rheumatic connective tissue disease.^1,2^ Clinically it is a heterogenous condition that ranges from a chronic disease that can remain stable over decades to a life-threatening condition. Disease manifestations can include vasculopathic complications such as digital ulcers, scleroderma renal crisis (SRC) and pulmonary arterial hypertension (PAH), as well as fibrotic complications such as skin fibrosis and interstitial lung disease (ILD).^1,3^ Anti-nuclear antibodies (ANA) are found in as many as 95% of patients with SSc.^4^ Many patients present with specific anti-centromeres antibodies (ACA), antibodies against topoisomerase 1 (Topo-1, also known as Scl-70) and/or RNA polymerase III (RP3) antibodies.^5^ The mechanisms underlying the development of these distinct antibodies in SSc are widely unclear, but accumulating data suggest a specific genetic background in combination with environmental and stochastic factors, as well as properties of the antigens themselves are key in antigen selection and antibody production.^6–9^ Moreover, the pathogenetic role of these autoantibodies in SSc is still a subject of ongoing research.^10^ Newer data suggest that antibodies could be pathogenic or at least contribute to the perennation of the disease.^8,11,12^ In addition, they have been established as strong predictors of disease outcome, of certain organ complications and therapeutic response.^13–15^ For example, Topo-1 and RP3 are more specific for dcSSc than ACA. In addition, ACA is associated with PAH without fibrosis while Topo-1 is frequently found in SSc patients with ILD.^6^ Moreover, RP3 autoantibodies are strongly associated with the incidence of scleroderma renal crisis and malignancy. Interestingly, immunochemistry analysis of cancer tissue from anti-RNAP-positive patients revealed a strong RNAP3 staining.^16^ These data support the idea that cancer initiates a specific immune response which, however, contributes aberrantly to the pathogenesis of SSc with a specific phenotype.^17^

Beside these typical associations, recently attention has been drawn to a phenomenon described as “inverted phenotype”, i.e, when there is a discordance between autoantibody and type of skin involvement, for example when a patient with anti-Topo-1 autoantibodies presents with limited skin disease.^18^ This group has been described as taking an intermediate risk position in terms of organ complications.^18–20^

There are also less frequently detected serum autoantibodies that are known to be associated with SSc, such as antibodies against TRIM-21/Ro 52, NOR-90, PM/Scl-75, PM/Scl-100, Th/To, Ku, fibrillarin, and PDGFR. Their clinical associations and frequencies are less well defined, as is the significance of positivity for multiple autoantibodies.^21^ In addition to classical autoantibodies, which are important in the diagnosis of SSc, functional autoantibodies to angiotensin II-type1- and endothelin-type-A-receptors have also been described for SSc.^22–25^ Via G protein-coupled receptor stimulation, these functional autoantibodies may be potentially pathogenic and responsible for different clinical manifestations.^22,26^ However, the associations with different SSc manifestations remain in parts controversial.^26,27^

As common for autoimmune diseases, SSc often coincides with other autoimmune diseases as Hashimoto thyroiditis or primary biliary cholangitis (PBC). PBC is a chronic cholestatic liver disease characterized by destruction of small intrahepatic bile ducts, leading to liver fibrosis and potential cirrhosis through resulting complications. The serological hallmark of primary biliary cirrhosis is the antimitochondrial antibody (AMA).^28^ The prevalence of clinically significant PBC in patients with systemic sclerosis is estimated to be 2.5%^29,30^, and up to 25% of SSc patients are positive for AMA.^31^ Moreover a positive ACA is reported in 9 – 30% of PBC patients.^29,32,33^ Some reports suggest that PBC-SSc is associated with a more favorable prognosis of PBC whereas others report increased mortality due to SSc.^29^ In addition, several studies have concluded that patients with PBC may have an increased risk for cardiovascular complications.^34,35^ However, these results are controversial.^36–38^ Concerning SSc, the cardiovascular effects of SSc-PBC or AMA positivity have not yet been defined.

Screening for antibodies is usually performed through indirect immunofluorescence staining (IIF). However, additional techniques, such as enzyme-linked immunosorbent assay (ELISA), immunodiffusion and immunoprecipitation, are used to identify specific SSc autoantibodies.^13,39,40^ Although, these immuno-assays may differ in terms of test characteristics and validation of results is crucial.^41,42^

In the present study, we investigated an extended autoantibody serum status in SSc patients using two different immunological methods and correlated the immunological phenotype of SSc patients with the clinical phenotype using cluster analysis. Here, we provide novel insights into the frequencies and associations of the antibodies especially with focus on inverted phenotypes, rarer antibodies as well as atypical antibodies such as AMA-M2.

## 2. Material and Methods

### 2.1 Patients

SSc patients from our center at the Department of Rheumatology, Charité -Universitätsmedizin Berlin, Germany were recruited between April 2013 and October 2018. The study protocol was approved by the Charité -Universitätsmedizin Berlin Ethics Committee (EA1/179/17). Written informed consent was obtained from each patient. The study was conducted in accordance with the principles of the Declaration of Helsinki.

Clinical parameters including sex, age, cutaneous subsets,^43^ age at onset of Raynaud’s phenomenon and age at onset of first non–Raynaud’s phenomenon symptom, disease duration and organ involvement were recorded. Variables collected included smoking history, digital ulcers, digital gangrene, calcinosis, highest modified Rodnan Skin Score (mRSS), systemic hypertension, hyperlipidemia, diabetes mellitus, myocardial infarction, angina pectoris, stroke, transitory ischemic attack (TIA), periphery arterial disease (PAD), PBC, PAH, interstitial lung disease (ILD), scleroderma renal crisis (SRC), heart involvement and myositis. Laboratory parameters (C-reactive protein [CRP], neutrophile count, hemoglobin, N-terminal pro-B-type natriuretic peptide [NT-proBNP]) were quantified from peripheral blood during clinical routine. Lung function was assessed via spirometry. Diffusing capacity for carbon monoxide (DLCO) was measured using the single-breath method. PAH was defined as a mean pulmonary artery pressure of ≥ 25 mmHg and a pulmonary capillary wedge pressure of ≤ 15 mmHg on right-sided heart catheterization. ILD was defined as the presence of pulmonary fibrosis on high-resolution computed tomography scan evaluated by experienced radiologists where HR-CT was performed upon clinical suspicion.

### 2.2 Antibody analysis

Serum aliquots were stored at -80°C prior to analyses. Sera were analyzed using commercially available ELISA and line immunoblot assay (all from EUROIMMUN Medizinische Labordiagnostika AG, Lübeck, Germany) according to the manufacturer’s instructions. The EUROLINE Systemic sclerosis (Nucleoli) profile (IgG) contains 13 recombinant antigens: DNA-topoisomerase I (Scl□70), centromere proteins A & B (CENP□A and CENP□B), RNA polymerase III (subunits RP11 and RP155), fibrillarin, NOR□90, Th/To, PM-Scl□100, PM-Scl□75, Ku, platelet-derived growth factor receptors (PDGFR), and Ro-52. Samples were analyzed at a dilution of 1:101 for line immunoblot testing. Autoantibodies were detected using alkaline phosphatase–labeled anti□human IgG. The EUROLINE flatbed scanner provides semi-quantitative results. Readings obtained with a signal intensity of 0-5 (-) and 6-10 (borderline) were considered negative. Positive measuring range was categorized as 11-25 (+), 26-50 (++), and above (+++).

ELISA detecting antibodies against M2-3E antigens were used with samples diluted 1:101. For Scl-70 or centromeres a dilution of 1:201 was applied. The Anti-M2-3E ELISA serves the serological detection of autoantibodies against mitochondria (AMA), precisely against AMA M2, by an antigen mix of native M2 and a recombinant fusion protein. Anti-human-IgG HRP (EUROIMMUN) were used as the standard secondary antibody conjugate for all ELISA. Results were reported using a positive cut-off at ≥20 RU/ml. Anti-AT_1_R and Anti-ET_A_R antibody serum levels were measured by ELISA (CellTrend GmbH, Luckenwalde, Germany [since 2012, One Lambda, Inc., Canoga Park, CA]) as described^24^, analogously to anti-ET_B_ receptor autoantibody quantification performed earlier in a partially overlapping cohort.^44^

### 2.3 Statistical Analysis

In an explorative approach, twostep cluster analyses with automated selection of the numbers of clusters were conducted to identify patient clusters with specific antibody patterns, and to understand which antibodies determine those specific profiles. To investigate the underlying patterns in the dataset principal components analysis of the autoantibody scores was performed in the MEDA package (R Library FactoMineR) in Jamovi.^45–47^ Dimensions 1 and 2 were used for clustering of SSc patients into autoantibody-defined subgroups unsing K-means algorithm. The optimal number of clusters was determined using the elbow method. Clinical associations with these autoantibody clusters were explored using the v test function, which compares each group mean to the overall mean. Continuous data are shown as mean and standard deviation (SD) or median and interquartile range (IQR), categorical data as count and percentages. Depending on the distribution, for group comparisons of serum levels t-test or Mann-Whitney *U* test was performed, for categorical variables on autoantibody status the Chi-Square test or Fisher’s test. Chi-Square test or Fisher’s test was also performed for clinical associations of the less frequent autoantibodies. *P*-values <0.05 were considered statistically significant. Due to the exploratory nature of the study no adjustment for multiple testing was applied. For statistical analysis, IBM SPSS Statistics version 28.0 and Jamovi^45^ were used.

## 3. Results

### 3.1 Demographic data

372 patients fulfilled the ACR/EULAR 2013 classification criteria for SSc. 238 (64.0%) were defined as lcSSc, 104 (28.0%) dcSSc and 30 (8.0) as systemic sclerosis sine scleroderma. Clinical data are summarized in **Table 1**. 87% of patients had Raynauds phenomenon. There was a prevalence of 34.9% of interstitial lung disease (ILD) when HR-CT was performed upon clinical suspicion and 12.4% of PAH when right heart catheter was performed upon clinical suspicion. Cardiac involvement was present in 8% of all patients, 35.8% had a history of digital ulcers (DU). Scleroderma renal crisis (SRC) was present in 2.7%. Arthritis was present in 9% of the patients, 8% had a history of calcinosis, and 7.8% had a history of malignancy.

**Table 1.** Demographic, clinical, and serologic characteristics (n=372)

|  |  |
| --- | --- |
| **Sex** |  |
| No. (%) female | 308 (82.8) |
| No. (%) male | 64 (17.2) |
| Female-to-male ratio | 5:1 |
| **Age, mean ± SD – years** |  |
| At onset RP | 45.79 ± 16.85 |
| At diagnosis | 47.99 ± 14.42 |
| **Disease duration, median (IQR) – years** |  |
|  | 7.45 (9.15) |
| **Disease classification, n (%)** |  |
| Diffuse (dcSSc) | 104 (28.0) |
| Limited (lcSSc) | 238 (64.0) |
| sine | 30 (8.0) |
| **Antinuclear antibody (ANA) positive, no. (%)** |  |
|  | 342 (95.3) |
| **SSc manifestations, n (%)** |  |
| Raynaud symptoms | 322 (86.6) |
| ILD | 130 (34.9) |
| PAH | 46 (12.4) |
| DU | 133 (35.8) |
| Calcinosis | 29 (8) |
| Cardiac involvement | 8 (2) |
| Arthritis | 33 (8.9) |
| SRC | 10 (2.7) |
| Myositis | 13 (3) |
| Terminal Organ failure | 9 (2.4) |
| Malignancy | 29 (7.8) |
| **Laboratory values, median (IQR)** |  |
| NT-proBNP – ng/L | 128.00 (216.00) |
| CRP – mg/dl | 0.75 (2.60) |
| Hb – mg/dl | 12.90 (2.30) |
| Neutrophil granulocytes | 4.97 (2.77) |
| **Cardiopulmonary parameters, mean ± SD** |  |
| FVC – %pred | 90.0 ± 20.3 |
| FEV1 - %pred | 82.8 ± 28.7 |
| DLCO - %pred | 52.9 ± 22.5 |
| LVEF - % | 56.8 ± 17.0 |

Disease duration refers to the time since first non-Raynaud symptom.
CRP, C-reactive protein; dcSSc, diffuse cutaneous SSc; DLCO, diffusing capacity for carbon monoxide; FEV1, forced expiratory volume per second; FVC, forced vital capacity; Hb, hemoglobin; ILD, interstitial lung disease; IQR, interquartile range; L, liter; lcSSc, limited cutaneous SSc; LVEF, left ventricular ejection fraction; n, number; NT-proBNP, N-terminal-pro-brain natriuretic peptide; PAH, pulmonary arterial hypertension; RP, Raynaud's phenomenon; SRC, scleroderma renal crisis; SSc, Systemic Sclerosis; %pred, percent predicted

### 3.2 Detected Autoantibodies and Coincidence of SSc-antibodies by Line Blot

Counts of individual autoantibodies and their expression, either monospecific or the number of times they appeared in combination with other autoantibodies, are shown in **Table 2**. In 39 patients (10.5%) no specific autoantibody could be detected by line blot analysis. However, of these 66.7% were positive for ANA. 125 patients (32.8%) had a monospecific autoantibody, while 138 (36.2%), 61 (16.0%), and 9 (2.4%) patients were positive for 2, 3, or more autoantibodies, respectively. The majority of patients were positive for ACA (ACA-CA or CB), Topo-1 or Ro52.

**Table 2.** Numbers and combinations of autoantibodies identified in the 372 SSc patients

|  | Ro-52 | PDGFR | Ku | PM75 | PM100 | Th/To | NOR90 | Fib | RP155 | RP11 | ACA-CB | ACA-CA | Topo-1 |
| --- | --- | --- | --- | --- | --- | --- | --- | --- | --- | --- | --- | --- | --- |
| Ro-52 |  | 0 | 5 | 4 | 8 | 4 | 5 | 0 | 4 | 5 | 39 | 47 | 28 |
| PDGFR |  |  | 0 | 0 | 0 | 0 | 0 | 0 | 0 | 0 | 0 | 0 | 0 |
| Ku |  |  |  | 1 | 2 | 0 | 1 | 0 | 0 | 0 | 3 | 3 | 6 |
| PM75 |  |  |  |  | 8 | 3 | 0 | 0 | 1 | 1 | 3 | 1 | 1 |
| PM100 |  |  |  |  |  | 4 | 0 | 0 | 0 | 1 | 4 | 3 | 5 |
| Th/To |  |  |  |  |  |  | 0 | 0 | 0 | 0 | 3 | 2 | 4 |
| NOR90 |  |  |  |  |  |  |  | 0 | 0 | 0 | 2 | 3 | 6 |
| Fib |  |  |  |  |  |  |  |  | 0 | 1 | 0 | 0 | 1 |
| RP155 |  |  |  |  |  |  |  |  |  | 12 | 1 | 1 | 1 |
| RP11 |  |  |  |  |  |  |  |  |  |  | 2 | 2 | 2 |
| ACA-CB |  |  |  |  |  |  |  |  |  |  |  | 127 | 2 |
| ACA-CA |  |  |  |  |  |  |  |  |  |  |  |  | 2 |
| Topo-1 |  |  |  |  |  |  |  |  |  |  |  |  |  |
| No. of mono-positive patients | 21 | 0 | 2 | 2 | 6 | 2 | 2 | 2 | 2 | 0 | 3 | 0 | 82 |
| Total No. of positive patients | 108 | 0 | 13 | 15 | 27 | 14 | 11 | 4 | 16 | 17 | 133 | 132 | 131 |

ACA, Anti-centromere antibody; Fib, Fibrillarlin; No., number; PDGFR, platelet-derived growth factor receptor; RP11 and 155; RNA polymerase III, subunit 11 and 155; Topo-1, anti-Topoisomerase-1 antibody

82 patients (22%) were monospecific for Topo-1 and 21 patients (5.6%) for Ro52. Co-expression of autoantibodies was common (**Tab. 2 and Suppl. Fig. 1**). Ro52 was the most frequent autoantibody that occurred in combination with other autoantibodies. Topo-1 was detected in 131 patients and ACA (CA and CB) were observed in 138 patients. 132 patients were ACA-CA and 133 were ACA-CB positive. 21 patients were found positive for RP3 (RP11 and RP155). PDGFR was not found in any of our patients. Co-expression of ACA and Topo-1 occurred rarely (4 of 372), as well as co-expression of Topo-1 and RP3 (3 of 372), and co-expression of ACA and RP3 (3 of 372).

### 3.3 Detection of Classical SSc Autoantibodies -ELISA vs. Line Blot

ACA and Topo-1 were measured both by ELISA and line blot. 132 patients were positive for anti-centromere antibodies in the ELISA and 138 patients were positive for ACA in the line blot (blot sensitivity for ACA 100%, blot specificity for ACA 97.5%). 125 patients were Topo-1 positive in the ELISA and 131 patients were Topo-1 positive in the line blot (blot sensitivity for Topo-1 100%, blot specificity for Topo-1 97.6%). Of the four patients double-positive for ACA and Topo-1 in the line blot, only one patient was also double-positive in the ELISA, three patients were ELISA-positive for Topo I only and one patient was both ACA and Topo-1 negative in the ELISA.

### 3.4 Cluster analysis

Principal component analysis (PCA) is an unsupervised machine learning technique which is applied to reduce the dimensionality of the input data, thus provides valuable insights even in very complex multivariate data sets. PCA of autoantibody levels was performed to examine underlying patterns in the clinical data set. It revealed strong negative associations between the 3 major autoantibodies (**Fig. 1**). Subsequently, K-means method was used to identify clusters in the data set. The K-means algorithm is an iterative method divides the data set into unique, non-overlapping subgroups (clusters), where each data point belongs to only one group. Using K-means clustering algorithm we were able to identify 4 autoantibody clusters, which we named after the assigned antibodies: RP3, Topo-1, Others and ACA (**Fig. 2**).

**Figure 1:**
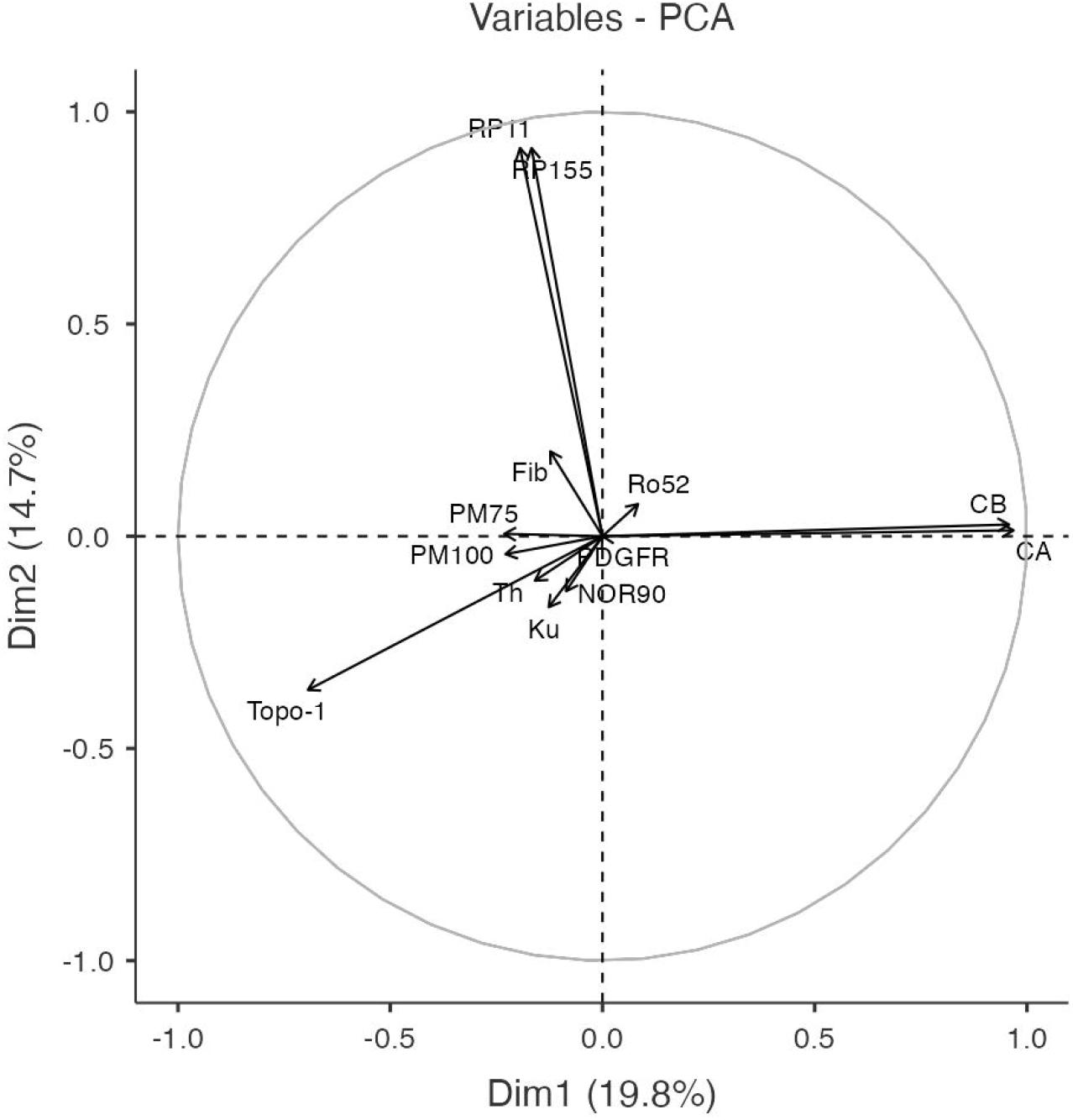
Correlation circle plot of the first 2 dimensions (Dim 1 and Dim 2) of the principal components analysis, which accounted for 34.5% of the total variance. This plot illustrates strong correlations between RNA polymerase 3 (RP3) epitope 11 and epitope 155 as well as between ACA epitopes A and B.

**Figure 2:**
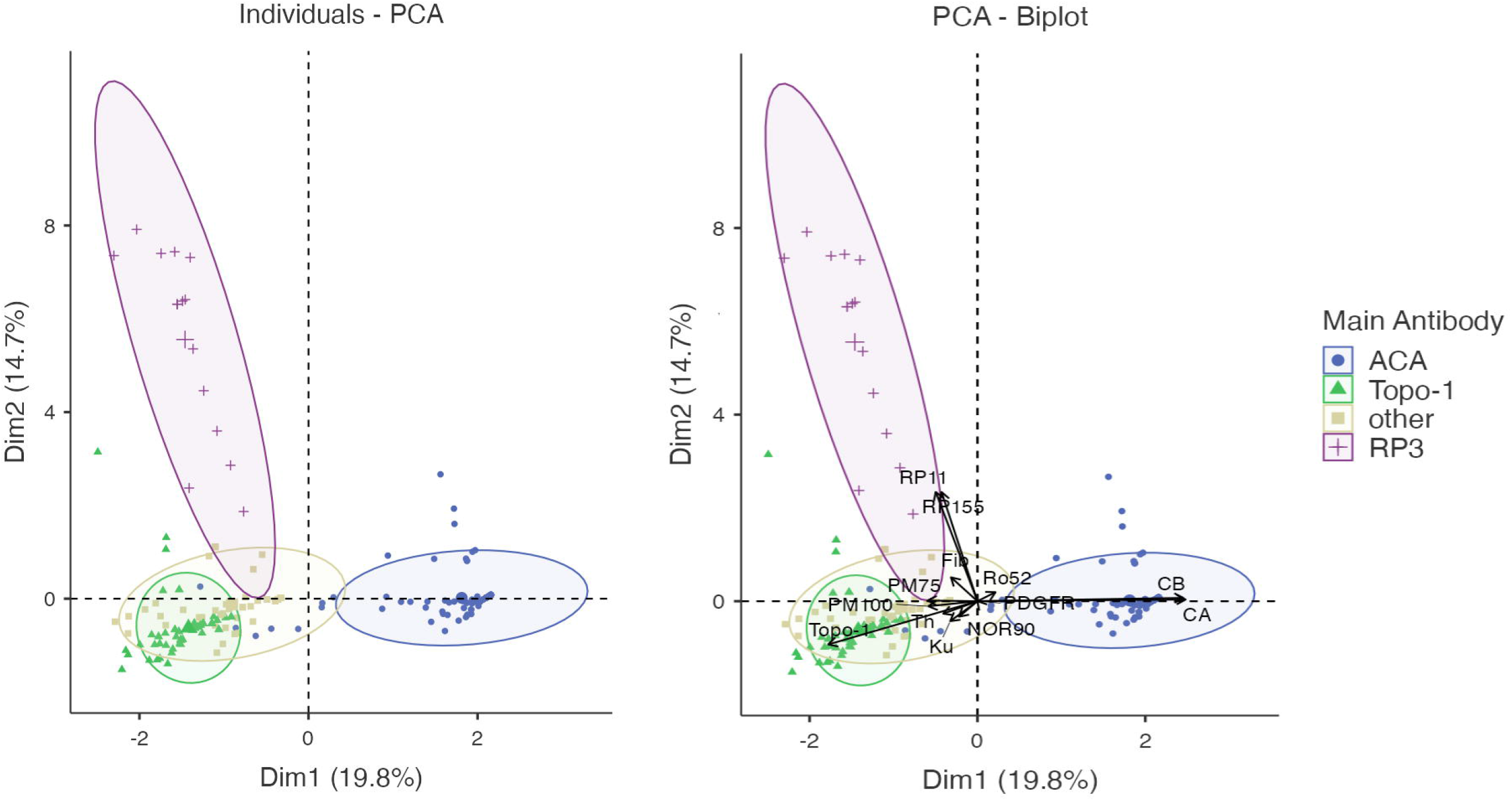
Clustering by K-means method revealed 4 autoantibody clusters: ACA, Topo-1, Others and RP3.

#### 3.4.1 Clinical associations of groups identified by cluster analysis

The 4 SSc clusters identified by principal components analysis were assessed with regard to their clinical characteristics. The results are shown in **Table 3**. Cluster *ACA* revealed many features consistent with limited cutaneous SSc, including female sex, lower neutrophil granulocyte count, PAD and coexisting PBC and was inversely associated with ILD, cardiac involvement, and impairment in lung function parameters. Cluster *RP3* demonstrated multiple features of diffuse cutaneous SSc, including high mRSS, elevated NT-pro BNP, and SRC. *Cluster Topo-1* also shows several features of dcSSc, including ILD, DU, elevated mRSS, reduced FVC, cases of terminal organ failure, and elevated neutrophil granulocyte count. Cluster *Others* was positively associated with older age of RP onset, myositis, reduced FEV1 and DLCO and elevated NT-pro BNP and inversely associated lcSSc and DU.

**Table 3.** Clinical manifestations of SSc clusters

| Demographic or clinical variable | All SSc patients<br>(n = 372) | Cluster |  |  |  | P value |
| --- | --- | --- | --- | --- | --- | --- |
|  |  | ACA<br>(n = 138) | Topo-1<br>(n = 127) | Others<br>(n = 53) | RP3<br>(n = 15) |  |
| Age at onset RP | 45.79 ± 16.99 | 45.76 ± 16.64 | 43.39 ± 16.13 † | 58.66 ± 17.10 † | 52.00 ± NA | 0.249 |
| Age at diagnosis | 47.99 ± 14.42 | 51.98 ± 14.81 † | 46.30 ± 14.49 ‡ | 45.69 ± 9.99 | 51.50 ± 14.99 | 0.172 |
| Disease duration * | 7.45 (9.15) | 7.00 (9.01) | 6.95 (9.05) | 7.10 (9.90) | 7.65 (3.68) | 0.989 |
| Female | 308 (82.8%) | 123 (89.1) ‡ | 98 (77.2) ‡ | 45 (84.9) | 10 (66.7) | **0.022** ‡ |
| lcSSc | 238 (64.0) | 120 (87.0) † | 58 (45.7) † | 27 (50.9) ‡ | 4 (26.7) ‡ | **<0.001** |
| dcSSc | 104 (28.0) | 11 (8.0) † | 64 (50.4) † | 14 (26.4) | 10 (66.7) ‡ | **<0.001** |
| Sine | 26 (7.0) | 7 (5.1) | 5 (3.9) | 12 (22.6) † | 1 (6.7) | **<0.001** |
| ILD | 130 (34.9%) | 20 (14.5) † | 74 (58.3) † | 19 (35.8) | 7 (46.7) | **<0.001** |
| Alveolitis | 28 (7.5%) | 2 (1.4) † | 19 (15.0) † | 7 (13.2) | 1 (6.7) | **<0.001** |
| PAH | 46 (12.4%) | 20 (14.5) | 16 (12.6) | 7 (13.2) | 1 (6.7) | 0.850 |
| DU | 133 (35.8%) | 44 (31.9) | 59 (46.5) ‡ | 13 (24.5) ‡ | 7 (46.7) | **0.014** ‡ |
| SRC | 10 (2.7%) | 2 (1.4) | 3 (2.4) | 1 (1.9) | 4 (26.7) † | **<0.001** |
| Myositis | 14 (3.8%) | 3 (2.2) | 3 (2.4) | 7 (13.2) ‡ | 1 (6.7) | **0.004** ‡ |
| Cardiac involvement | 10 (2.7%) | 1 (0.7) ‡ | 6 (4.7) | 2 (3.8) | 1 (6.7) | 0.208 |
| Arthritis | 33 (8.9%) | 9 (6.5) | 13 (10.2) | 4 (7.5) | 2 (13.3) | 0.634 |
| mRSS | 6.63 ± 8.09 | 4.70 ± 6.18 † | 9.31 ± 8.89 † | 5.31 ± 7.86 | 11.50 ± 11.69 ‡ | **<0.001** |
| Calcinosis | 33 (8.9%) | 18 (13.0) | 9 (7.1) | 5 (9.4) | 0 (0.0) | 0.221 |
| Vasculitis | 5 (1.3%) | 1 (0.7) | 2 (1.6) | 1 (1.9) | 1 (6.7) | 0.344 |
| Malignancy | 29 (7.8%) | 9 (6.5) | 9 (7.1) | 3 (5.7) | 3 (20.0) | 0.266 |
| LVEF - % | 56.8 ± 16.9 | 56.0 ± 17.6 | 58.7 ± 12.5 | 58.9 ± 17.3 | 57.5 ± 15.1 | 0.716 |
| FVC - %pred | 90.9 ± 20.3 | 95.9 ± 18.6 † | 84.8 ± 20.1 † | 86.3 ± 23.0 | 92.0 ± 11.5 | **<0.001** |
| FEV1 - %pred | 82.8 ± 28.7 | 88.6 ± 27.0 ‡ | 83.5 ± 26.0 | 70.7 ± 33.8 † | 86.5 ± 15.0 | **0.009** ‡ |
| DLCO - %pred | 52.9 ± 22.5 | 58.5 ± 21.0 † | 51.5 ± 21.0 | 45.4 ± 23.0 ‡ | 50.6 ± 19.0 | **0.016** ‡ |
| Term. organ failure | 9 (2.4%) | 2 (1.4) | 6 (4.7) ‡ | 0 (0.0) | 0 (0.0) | 0.163 |
| NT-proBNP * | 128.00 (216.00) | 115.00 (291.50) ‡ | 149.00 (197.00) | 134.00 (142.00) ‡ | 179.00 (1381.00) ‡ | 0.108 |
| CRP * | 0.75 (2.60) | 0.75 (2.43) | 1.00 (2.88) | 0.50 (2.22) | 0.90 (0.40) | 0.989 |
| Hb | 11.30 ± 4.80 | 11.16 ± 4.83 | 11.90 ± 4.25 | 11.26 ± 5.03 | 12.52 ± 1.06 | 0.583 |
| Neutrophile | 5.38 ± 2.65 | 4.73 ± 2.06 † | 5.77 ± 3.04 ‡ | 5.68 ± 2.98 | 5.12 ± 2.24 | 0.167 |
| History of smoking | 43 (11.6%) | 19 (13.8) | 9 (7.1) | 7 (13.2) | 1 (6.7) | 0.150 |
| Hyperlipidemia | 16 (4.3%) | 8 (5.8) | 5 (3.9) | 2 (3.8) | 0 (0.0) | 0.704 |
| Diabetes | 12 (3.2%) | 4 (2.9) | 2 (1.6) | 4 (7.5) | 0 (0.0) | 0.163 |
| Myocardial infarction | 6 (1.6%) | 3 (2.2) | 1 (0.8) | 2 (3.8) | 0 (0.0) | 0.515 |
| Stroke | 9 (2.4%) | 2 (1.4) | 4 (3.1) | 2 (3.8) | 0 (0.0) | 0.652 |
| Angina pectoris | 14 (3.8%) | 7 (5.1) | 2 (1.6) | 2 (3.8) | 0 (0.0) | 0.378 |
| TIA | 3 (0.8%) | 1 (0.7) | 0 (0.0) | 0 (0.0) | 0 (0.0) | 0.701 |
| PAD | 13 (3.5%) | 9 (6.5) ‡ | 1 (0.8) | 0 (0.0) | 0 (0.0) | **0.018** ‡ |
| MI/TIA/Stroke/pAVK | 35 (9.4%) | 18 (13.0) | 6 (4.7) ‡ | 5 (9.4) | 0 (0.0) | 0.099 |
| PBC | 17 (4.6%) | 14 (10.1) † | 1 (0.8) ‡ | 2 (3.8) | 0 (0.0) | **0.004** ‡ |

Results are indicated as number (percentage) or mean ± standard deviation.
† P <0.001 versus the overall mean, as determined by v test.
‡ P <0.05 versus the overall mean, as determined by v test.
\* median ± IQR are shown
CRP, C-reactive protein; dcSSc, diffuse cutaneous SSc; DLCO, diffusing capacity for carbon monoxide; Du, digital ulcers; FEV1, forced expiratory volume per second; FVC, forced vital capacity; Hb, hemoglobin; ILD, interstitial lung disease; IQR, interquartile range; L, liter; lcSSc, limited cutaneous SSc; LVEF, left ventricular ejection fraction; MI, myocardial infarction; mRSS, modified Rodnan Skin Score; n, number; NT-proBNP, N-terminal-pro-brain natriuretic peptide; PAD, peripheral arterial disease; PAH, pulmonary arterial hypertension; PBC, primary biliary cholangitis; RP, Raynaud’s phenomenon; SRC, scleroderma renal crisis; SSc, Systemic Sclerosis; TIA, transient ischemic attack; %, percent, %pred, percent predicted

**Table 4:**
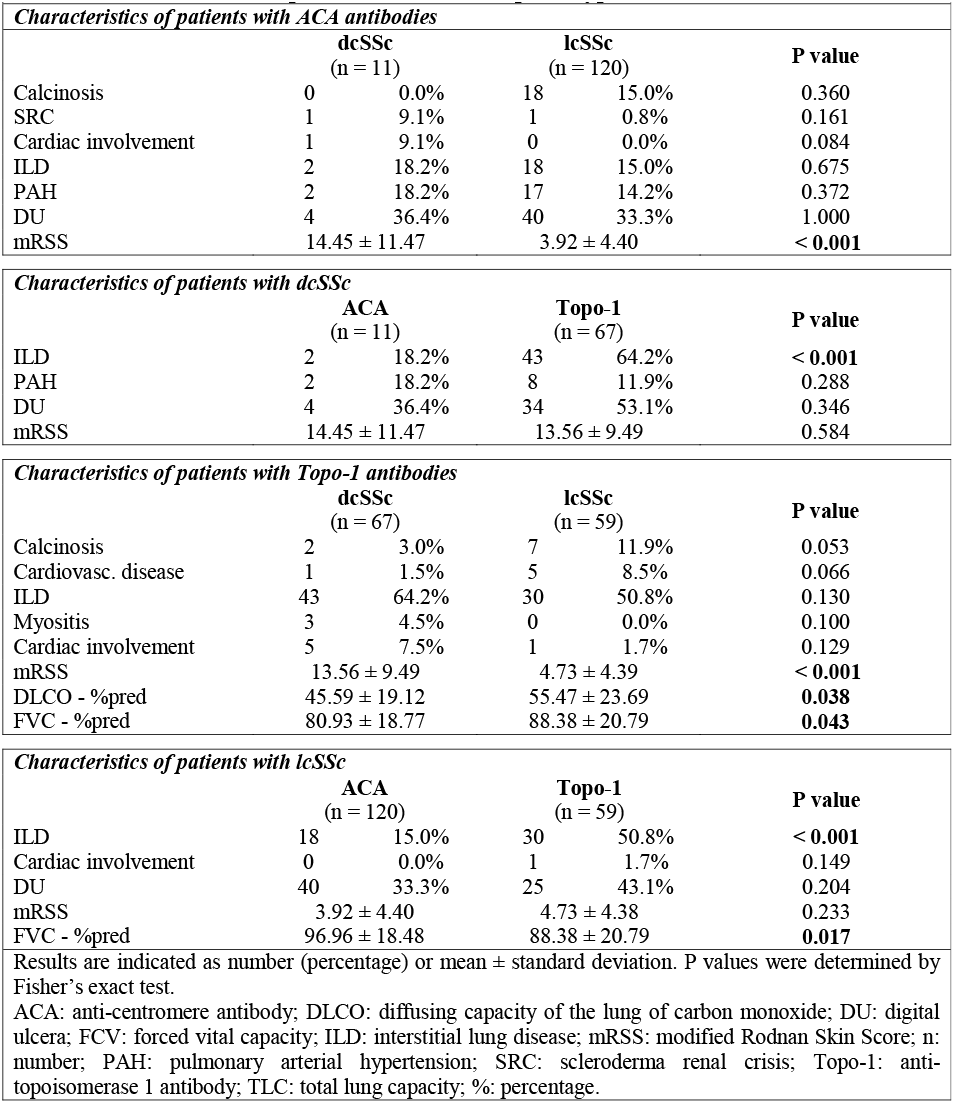
Characteristics of patients with inverted phenotype.

|  | **dcSSc**<br>(n = 11) |  | **lcSSc**<br>(n = 120) |  | **P value** |
| --- | --- | --- | --- | --- | --- |
| Calcinosis | 0 | 0.0% | 18 | 15.0% | 0.360 |
| SRC | 1 | 9.1% | 1 | 0.8% | 0.161 |
| Cardiac involvement | 1 | 9.1% | 0 | 0.0% | 0.084 |
| ILD | 2 | 18.2% | 18 | 15.0% | 0.675 |
| PAH | 2 | 18.2% | 17 | 14.2% | 0.372 |
| DU | 4 | 36.4% | 40 | 33.3% | 1.000 |
| mRSS | 14.45 ± 11.47 |  | 3.92 ± 4.40 |  | **< 0.001** |

***Characteristics of patients with dcSSc***
|  | **ACA**<br>(n = 11) |  | **Topo-1**<br>(n = 67) |  | **P value** |
| --- | --- | --- | --- | --- | --- |
| ILD | 2 | 18.2% | 43 | 64.2% | **< 0.001** |
| PAH | 2 | 18.2% | 8 | 11.9% | 0.288 |
| DU | 4 | 36.4% | 34 | 53.1% | 0.346 |
| mRSS | 14.45 ± 11.47 |  | 13.56 ± 9.49 |  | 0.584 |

***Characteristics of patients with Topo-1 antibodies***
|  | **dcSSc**<br>(n = 67) |  | **lcSSc**<br>(n = 59) |  | **P value** |
| --- | --- | --- | --- | --- | --- |
| Calcinosis | 2 | 3.0% | 7 | 11.9% | 0.053 |
| Cardiovasc. disease | 1 | 1.5% | 5 | 8.5% | 0.066 |
| ILD | 43 | 64.2% | 30 | 50.8% | 0.130 |
| Myositis | 3 | 4.5% | 0 | 0.0% | 0.100 |
| Cardiac involvement | 5 | 7.5% | 1 | 1.7% | 0.129 |
| mRSS | 13.56 ± 9.49 |  | 4.73 ± 4.39 |  | **< 0.001** |
| DLCO - %pred | 45.59 ± 19.12 |  | 55.47 ± 23.69 |  | **0.038** |
| FVC - %pred | 80.93 ± 18.77 |  | 88.38 ± 20.79 |  | **0.043** |

***Characteristics of patients with lcSSc***
|  | **ACA**<br>(n = 120) |  | **Topo-1**<br>(n = 59) |  | **P value** |
| --- | --- | --- | --- | --- | --- |
| ILD | 18 | 15.0% | 30 | 50.8% | **< 0.001** |
| Cardiac involvement | 0 | 0.0% | 1 | 1.7% | 0.149 |
| DU | 40 | 33.3% | 25 | 43.1% | 0.204 |
| mRSS | 3.92 ± 4.40 |  | 4.73 ± 4.38 |  | 0.233 |
| FVC - %pred | 96.96 ± 18.48 |  | 88.38 ± 20.79 |  | **0.017** |

Results are indicated as number (percentage) or mean ± standard deviation. P values were determined by Fisher's exact test.
ACA: anti-centromere antibody; DLCO: diffusing capacity of the lung of carbon monoxide; DU: digital ulcers; FVC: forced vital capacity; ILD: interstitial lung disease; mRSS: modified Rodnan Skin Score; n: number; PAH: pulmonary arterial hypertension; SRC: scleroderma renal crisis; Topo-1: anti-topoisomerase 1 antibody; TLC: total lung capacity; %: percentage.

#### 3.4.2 ACA and Topo-1 with inverted phenotype

59 Topo-1 positive patients (45.0%) had lcSSc and 11 ACA positive patients had dcSSc (**Tab. 4**). Compared to Topo-1 dcSSc patients, Topo-1 lcSSc patients had a higher DLCO and FVC and less skin sclerosis. Moreover, they tended to have a higher prevalence of calcinosis, an increased risk for cardiovascular disease and a lower prevalence of ILD and less risk for myositis. Compared to ACA lcSSc patients, ILD was more prevalent in Topo-1 lcSSc than in ACA lcSSc patients and FVC was significantly decreased in Topo-1 lcSSc, compared with ACA lcSSc. Also, ILD was slightly less prevalent in the Topo-1 lcSSc than in Topo-1 dcSSc patients. Compared to ACA lcSSc patients, ACA dcSSc patients tended to develop calcinosis cutis less frequently. Compared to Topo-1 dcSSc, ACA dcSSc patients were less likely to suffer from ILD.

#### 3.4.3 Other SSc-associated autoantibodies

Univariate analysis was performed to investigate clinical associations with the presence of TRIM-21/Ro52, NOR-90, PM/Scl-75, PM/Scl-100, Th/To, Ku, fibrillarin (fib, also known as U3RNP) and PDGFR. Statistically significant associations or those approaching statistical significance are summarized in **Table 5**.

**Table 5.**
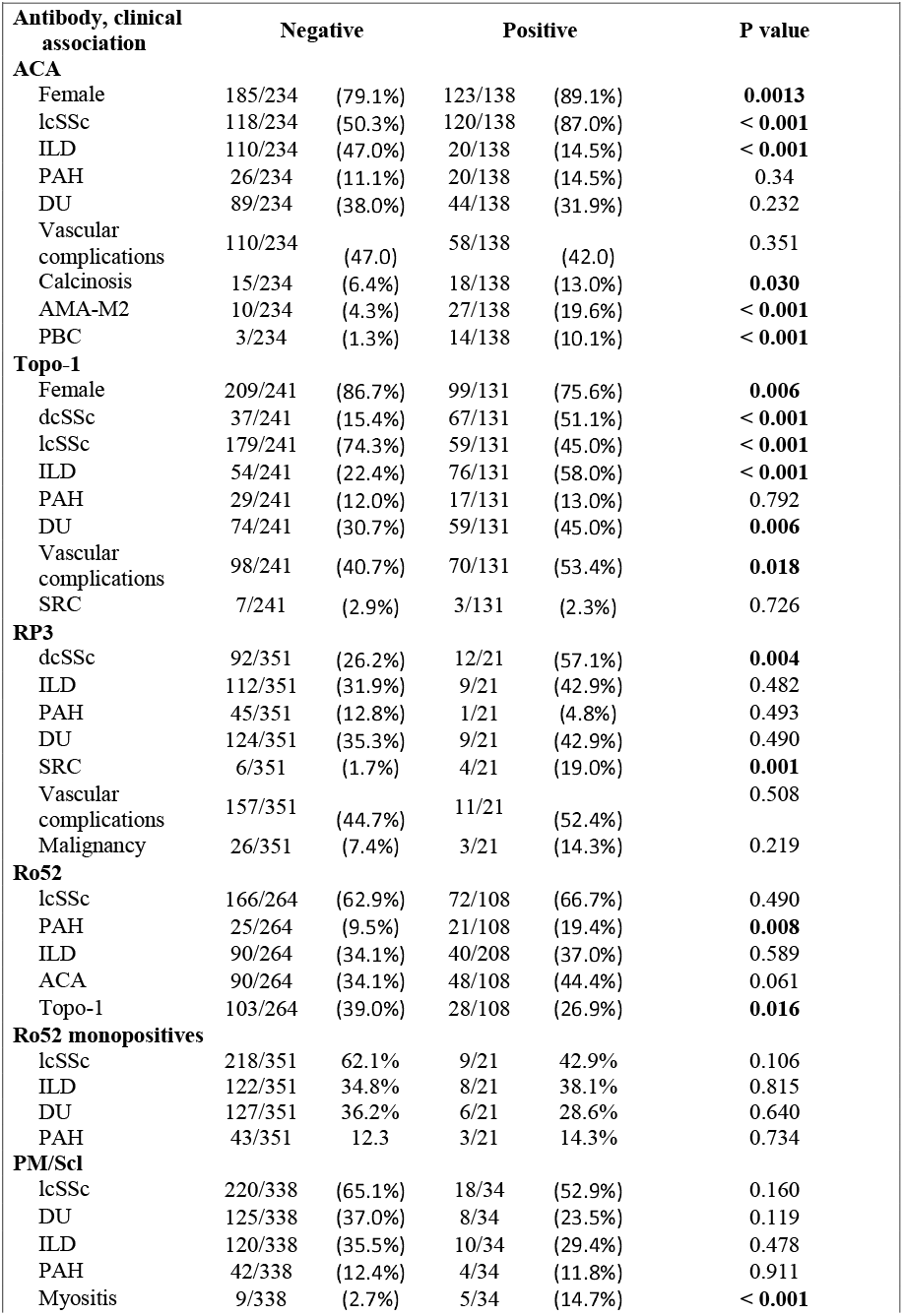
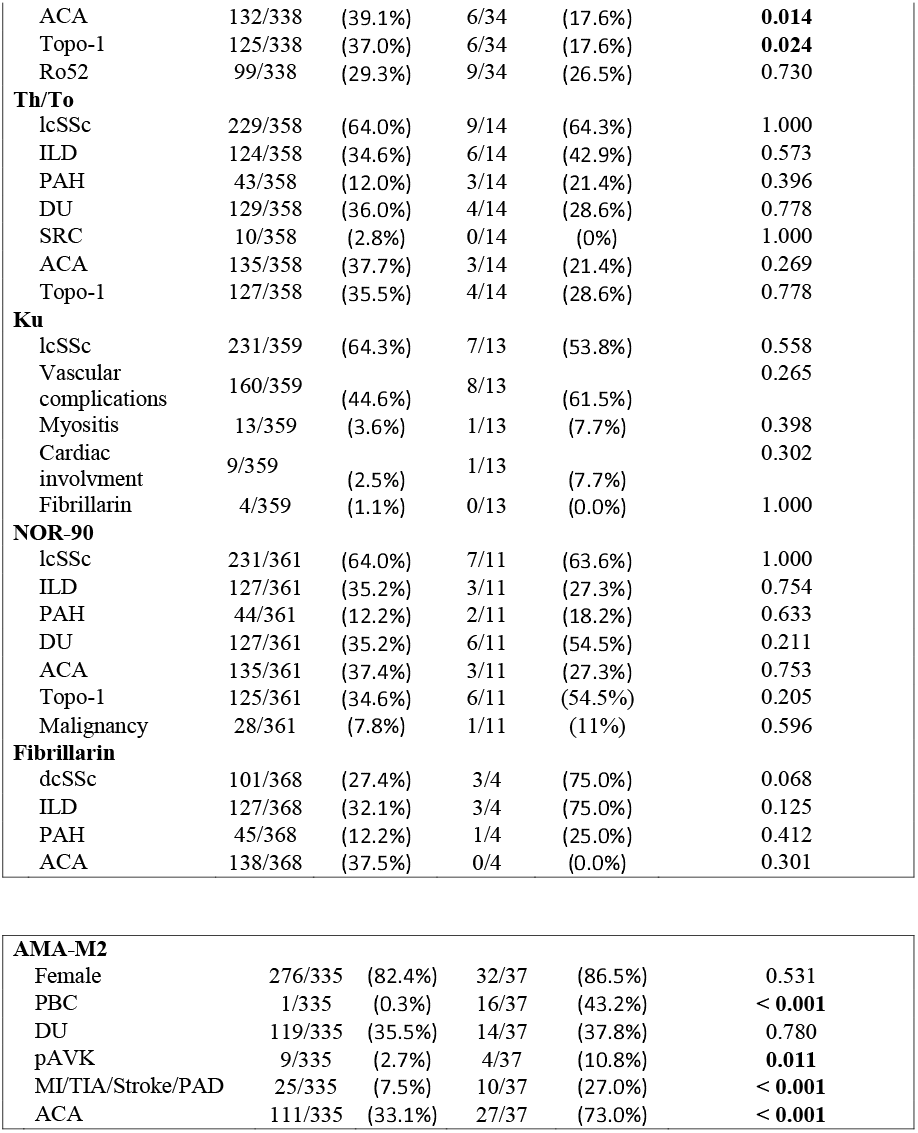
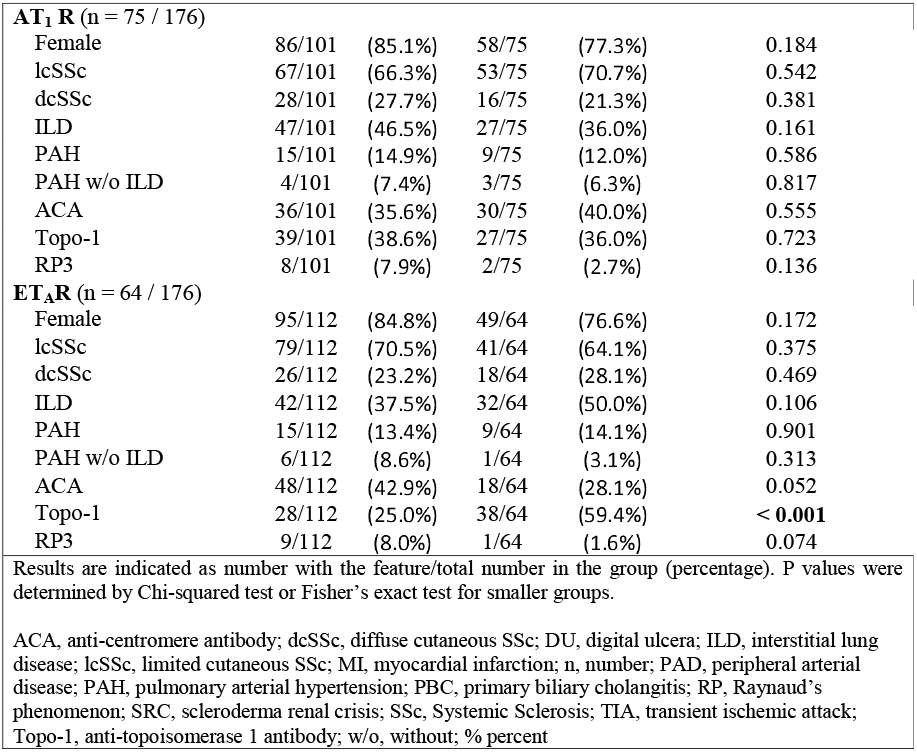
Clinical characteristics of the SSc-associated autoantibodies

Ro52 and PM/Scl-75 or PM/Scl-100 were the most common other SSc-associated autoantibodies. 108 patients were positive for Ro52. Ro52 was more commonly seen in the presence of ACA (48/108). Only 26.9% of the Ro52-positive patients were Topo-1 positive. 19.4% of the Ro52 positive patients were diagnosed with PAH, compared to 9.5% of Ro52 negative patients (*p* = 0.008). 32.40% of Ro52 positive patients were diagnosed with ILD, compared with 32.6% of the patients negative for Ro52. 21 patients (5.6%) were monospecifically positive for Ro52. However, monospecificity for Ro52 was not associated with a distinct clinical disease pattern.

Given the low numbers of patients with antibodies against PM/Scl-75/100 (34 of 372), Th/To (24 of 372), Ku (13 of 372), fibrillarin (4 of 372) and NOR-90 (11 of 372), any associations reported should be interpreted with caution: In our cohort, 34 patients were positive for PM/Scl-75/100 (27 female, 7 male), and 12 were monospecifically positive for PM/Scl-75/100. PM/Scl was associated with myositis and inversely associated with coexisting ACA or Topo-1. In addition, there was a negative trend for DU. Ku was found in 13 patients equally in lcSSc and dcSSc. All patients had Raynaud’s phenomenon. However, no significant association with a distinct clinical pattern was found. Fib positve patients had diffuse cutaneous involvement and a tended to have ILD. 21.4% (n=3) of Th/To positive patients had PAH. Patients with NOR90 tended to show an increased incidence of DU and a coexistence of Topo-1.

### 3.5 Clinical characterisics of AMA-M2 positive SSc patients

In our cohort, 37 patients (9.9%) were positive for AMA-M2 (86% female) (**Tab. 5**). 73.0% patients were positive for ACA (p < 0.001), 5 patients were Topo-1 positive and 2 patients were positive for RP3. 16/37 AMA-M2 positive patients were diagnosed with PBC (p < 0.001). AMA-M2 positive patients with diagnosed PBC tended to have higher γ-glutamyltransferase (gGT), compared with AMA-M2 positive patients without a diagnosis of PBC (87.1 ± 98.8 vs. 43.7 ± 41.1; p = 0.076). 10.8% of AMA-M2 positive patients had PAD compared with 2.7% of AMA-M2 negative patients (p = 0.01) and 27.0% of AMA-M2 positive patients had cardiovascular disease (such as mycadial infarction, TIA, stroke or PAD), compared to 7.5% of AMA-M2 negative patients (p < 0.001). The increased cardiovascular risk applies both to AMA-M2 positive patients with and without PBC (p = 0.004 and p = 0.02).

### 3.6 Vascular receptor autoantibodies

Anti-AT_1_R and anti-ET_A_R autoantibodies were assessed in 176 SSc patients. Demographic data, clinical manifestations, disease duration and serology are shown in **suppl. Tab. 1**. As described before, cut-off values were 9.2 units for anti-AT_1_R and 10.4 units for anti-ET_A_R antibodies.^25^ 42.6% of patients were positive for antibodies against AT_1_R and 36.4% were positive for anti-ET_A_R antibodies, respectively. A clear correlation between anti-AT_1_R and anti-ET_A_R auto-antibodies was present in patients with SSc (r = 0.75; **Suppl. Fig. 2A**) as described before.^25^ In our SSc cohort, we did not find significant association between positive AT1R or ETAR status in the serum status of SSc patients with the presence of ILD or PAH (**Tab. 4** and **Suppl. Fig. 2B and C**). Interestingly, patients with anti-ET_A_R autoantibodies were significantly more often Topo-1 positive compared with patients without anti-ET_A_R autoantibodies (**Tab. 5**).

## 4. Discussion

Autoantibodies are one of the strongest predictors of disease course, outcome and therapeutic response in patients with SSc.^4,15^ Nevertheless, uncertainties remain in the clinical association for some autoantibodies. In the present study, we were able to demonstrate that the classical SSc antibodies (ACA, Topo-1 and RP3) show strong negative correlations and form their own clusters to which certain clinical manifestations of SSc can be assigned.

Traditionally, SSc has been classified according to the extent of skin fibrosis.^43^ However, dermatologic findings are a dynamic process, so the early classification of SSc as limited may need to be revised as the disease progresses.^20,21,48^ In contrast, autoantibodies are a consistent feature of the disease, and it is rare for an autoantibody to disappear. Thus, recent studies have tackled the issue showing that dermatologic evaluation only is not sufficient to classify patients. In our present study, which was performed in a representative cohort of Caucasian SSc patients^49^, we could identify different clusters based on clinical features and autoantibody profiles, and thus confirm and further extend the results from others.^21,50^

Due to the nuclear nature of the targets of SSc specific antibodies, both the origin and the pathological role of these antibodies were unclear for a long time. Antibody generation includes the release of neoantigens, post-translational modifications and antigen presentation. In SSc, apoptotic blebs of endothelial cells and Topo-1 release from by apoptotic blebs are described.^8,51,52^ In addition, the release of neutrophil extracellular traps (NETs), also called NETosis, has also been highlighted in SLE as a contributor to autoimmunity and a source for autoantibodies. Recently, Didier et al. could demonstrate evidence for NETosis also in SSc.^53^ As previously reported, autoantibodies can also be induced as part of the immune response to malignancy.^54^ We were able to show in our cohort that patients in the RP3 cluster tend to have an increased prevalence of malignancies, which may be involved in the formation of these antibodies. For antigen presentation, major histocompatibility complex genes (human leukocyte antigen, HLA) are most relevant. In SSc, specific HLA-alleles may provide susceptibility to classical disease-specific autoantibodies. For example, Topo-1 was associated with DRB1^*^11:01/^*^11:04 in North American Caucasians, DPB1^*^13:01 in both African American and European-American patients.^7^ ACAs were found associated with DQB1^*^05:01/^*^26 alleles and RP3 with

DRB1^*^04:04, DRB1^*^11 and DQB1^*^03.^7^ These novel findings may explain why co-expression of multiple SSc-type antibodies is rare and why specific clinical clusters can be assigned to these antibodies.^6,55^ In most instances we were able to confirm these associations:

ACA is the most frequently seen autoantibody in SSc patients.^4^ In our cohort, 37% of patients were ACA positive (**Tab. 3**). In literature, the frequency of ACA in patients with SSc has been reported to be 20–30% overall, however, it varies among different ethnic populations.^13,56,57^ Mierau et al. found 35.9% of patients to be ACA positive.^49^ The detection of ACA led to its inclusion in the ACA cluster in our analysis. In line with previous reports, ACA was associated with lcSSc and inversely associated with ILD. Moreover, ACA positive patients tended to have more PAH and calcinosis. The association of ACA with pulmonary hypertension has been observed in several previous reports ^5,13,14,21,49,58–63^, but not all of them.^48,49^ In addition, 19.6% of ACA-positive SSc patients were positive for AMA-M2 and 10.1% were diagnosed with PBC.^64^

The prevalence of Topo-1 in our cohort (34%) is in line with the numbers published by others.^5,49,59^ The presence of the Topo-1 led to its assignment to the Topo-1 cluster in our cluster analysis. Topo-1 was significantly associated with dcSSc, however, 44.9% of Topo-1 positive patients had lcSSc. These numbers slightly exceed those reported in the literature.^65^ Here, again the dynamic nature of skin involvement should be noted, and it cannot be excluded that some of these patients may develop diffuse skin involvement in the course of their disease. The cluster Topo-1 was associated with a higher risk for pulmonary fibrosis and digital ulcers. These results are in line with previous findings.^21,49^ The value of Topo-1 as a predictor of renal crisis is uncertain and controversial in the literature.^21,49,60,66^ In our cohort, we did not see an association between Topo-1 and SRC.

The third antibody that led to assignment to a clinical cluster was RP3. In line with previous reports, the RP3 cluster in our cohort was associated with dcSSc and higher risk for renal crisis. ^67–70^

In their study, Patterson et al. found 2 different autoantibody clusters, depending on the intensity of R3 staining (cluster RNAP III strong and cluster RNAP III weak).^21^ Since RP3 titers may change over time^68^, the authors concluded, that the clusters RNAP III strong and RNAP III weak may represent different temporal stages of SSc disease.^21^ However, we could not find this distinction of the cluster in our analysis. It should be noted that Patterson and colleagues included 81 patients with RP3, whereas we only included 21.

Co-expression between the 3 major autoantibodies was only rarely observed in this cohort (**Tab. 2**) but it has been also reported in previous reports.^21,49,71–73^ However, presence occasionally may represent a false-positive result. In our cohort, four patients were positive for ACA and Topo-1 in the line blot but only one patient in the ELISA. From an immunological point of view and considering that different genetic predispositions may result in different antibodies, laboratory errors are a reasonable explanation.

When multiple autoantibodies were examined in SSc patients in this study, ACA, Topo I, and RP3 remained the autoantibodies with the most pronounced inverse correlation with each other and are each associated with characteristic clinical features. While co-expression of any of these 3 major autoantibodies remained rare (**Tab. 2**), co-expression with other autoantibodies has been found to be frequent.^21,73^

Despite the common associations between specific SSc antibodies and clinical characteristics (Topo-1 and dcSSc, ACA and lcSSc), inverted phenotypes of SSc which refers to the phenomenon that ACA positive patients show a dcSSc phenotype or Topo-1 positive patients show a lcSSc phenotype have been described.^20,74–78^ In our study, Topo-1 lcSSc patients had less lung involvement than Topo-1 dcSSc patients. However, Topo-1 lcSSc patients had an increased risk for ILD, when compared with ACA lcSSc (**Tab. 4**). This indicates that inverted phenotypes should be considered as a separate group occupying an intermediate risk position in terms of organ complications and underlines the importance of antibody detection.^20^ In this context, the faSScinate study, which investigated tocilizumab in patients with dcSSc, indicated that the autoantibody status is important for the therapy. Tocilizumab showed significant decrease in rates of lung function decline in Topo-1 positive patients but not in Topo-1 negative patients in phase 2 and 3 studies.^15,79–81^

The rarer SSc-associated autoantibodies were not found by principal components analysis to significantly contribute to subclassification. These patients were captured in cluster Other. This cluster was positive associated with higher risk of myositis and elevated NT-proBNP. Negative associations were found for lung function parameters (FVC and DLCO) and DU. Although, the rarer SSc-associated autoantibodies did not contribute to subclassification, clinical associations were assessed in this study (**Tab. 5**). In our cohort, Ro52 positivity was associated with presence of PAH. This association has also been discovered by others.^21,82 79,80^Others have reported an association between Ro52 and ILD.^83^ However, Patterson et al.^21^ or Lee et al.^82^ were not able to replicate these findings. In our study, we were neither able to find this association.

Anti-PM/Scl is reported to predict for lcSSc, myositis, calcinosis and no serious internal organ involvement with good prognosis.^49,84,85^ In our cohort, PM/Scl was associated with myositis, and by trend with lcSSc and DU. Moreover, PM/Scl was negatively associated with co-expression of both ACA and Topo-1. Autoantibodies to Th/To were reported to be specific for SSc or Raynaud’s disease with a short disease duration and lcSSc.^86^ Moreover, a high risk for severe organ involvement such as ILD and development of PAH and therefore a worse overall prognosis has been reported.^87,88^ However, we did not find an association with ILD. For anti-NOR90 antibodies an association with malignancy is described in literature.^89^ In our cohort, we did not find this association. Moreover, case reports suggests that NOR90 might be associated with lcSSc, mild internal organ involvement and a favorable prognosis.^90,91^ We can neither disprove nor confirm these observations. Anti-Ku has been reported in sera from patients with other connective tissue diseases, such as SLE and overlap syndrome, especially myositis.^92,93^ However, we could not prove this association with myositis in our cohort. Likewise, others could not find this correlation, neigher.^21^ Patients with anti-fibrillarin are reported to have dcSSc and vasculopathy, including DU and PAH.^4,94,95^ We can confirm the association to dcSSc in our very small cohort of anti-fibrillarin positive SSc patient. In addition, we see a trend toward increased risk for ILD. Baroni et al.^96^ reported autoantibodies against the platelet-derived growth factor receptor (PDGFR) in patients with SSc and for these autoantibodies a pathogenic role is suspected. ^97–99^ However, we did not find any patient with this antibody in our cohort.

AT_1_R- and ET_A_R-autoantibodies have been shown to be more frequently positive in patients with PAH secondary to SSc or other connective tissue disease when compared to patients with idiopathic PAH or chronic thromboembolic pulmonary hypertension.^25^ Moreover, their presence has been shown to be associated with vasculopathic changes like endothelial activation and smooth muscle contraction^23,25,100^ and their presence was associated with SSc-related manifestations like PAH, renal crisis, digital ulcers, lung fibrosis, and mortality.^24,25^ However, we were not able to confirm an association of AT_1_R- and/or ET_A_R-autoantibody positivity with PAH, renal crisis, digital ulcers or ILD. This might be due to the fact, that our cohort is significantly smaller than the cohort of Riemekasten et al.^24^. On the other hand, our findings are in line with findings by Ilgen et al. ^27^ and Bankamp et al. ^26^ who were also not able to find these associations.

The significance of a coexisting PBC with SSc is not well understood. To our knowledge, we are the first who demonstrated that SSc patients with positive AMA-M2 had a significantly higher risk of cardiovascular events. No significant difference was found between AMA-M2 positive patients diagnosed with PBC and those without PBC. Both patient groups have an increased cardiovascular risk. Interestingly, as in SSc, also in PBC endothelial dysfunction is thought to be involved in the pathophysiology^101^ and Fonollosa et al. found a high prevalence of nailfold capillary abnormalities characteristic of systemic sclerosis in patients with PBC.^102^

For the determination of antibodies in SSc, different methods exist, each of which has strengths and weaknesses.^103,104^ For clinical practice, simple and cost-efficient methods such as ELISA and immunoblotting have become established.^105^ In our study we used both methods for the determination of ACA and Topo-1 and could show a good agreement for the EUROLINE blot compared to ELISA. However, it should be noted that both methods are based on similar principles and thus share common sources of error, such as non-specific binding and cross-reactivity.^42,105^ Furthermore, it has to be considered that the other antibodies we determined and on which our cluster analysis is based were only determined with a single technique. Mierau et al. analysed sera of 863 SSc patients using different methods including IIF, a comparable line immunoassay as we did, immunoprecipitation and immunodiffusion.^49^ These authors found a high concordance between the results from those different techniques and broadly similar results to ours on the frequencies and clinical associations of the different antibodies.

## 5. Conclusion

In our study, we could confirm and extend previously described correlation between certain autoantibodies and clinical phenotype in SSc in a well characterized Caucasian cohort by an unsupervised machine learning technique. We used line blot data that was partially validated by confirming results by ELISA to determine SSc associated autoantibodies. Although several antibodies were included in our analysis, we found, that the dominant SSc antibody most strongly predicted the clinical phenotype. However, we could highlight the importance of analyzing an extended spectrum of autoantibodies, including antibodies to AMA-M2, which were associated with a prior unknown elevated cardiovascular risk, to best assess clinical phenotypes, as well as organ complications and comorbidities. Despite the described advantages of determining antibody profiles, clinical examination is not obsolete. Especially for inverted phenotypes, we were able to show, that these patients form a special subgroup and that the antibody status of the patients must be evaluated in combination with the clinical examination.

## Data Availability

All data produced in the present study are available upon reasonable request to the authors

## Acknowledgements

The authors would like to thank Daniel Grund and Udo Schneider for their support on patient recruitment. CT and ES are participants in the BIH-Charité Clinician Scientist Program funded by the Charité – Universitätsmedizin Berlin and the Berlin Institute of Health. The authors gratefully acknowledge Martina Lehmann and Christina Straßer for their excellent technical assistance and Dr. Alexander Kühnl for his valuable scientific communication.

## Ethical statement

The study protocol was approved by the ethics committee of Charité – Universitätsmedizin Berlin (EA1/179/17). A written informed consent was obtained from each patient.

## Author contributions

Study design: JH, ES, DH, GRB. Study conduct: JH, VC, CK, CT. Data collection: JH, VC, CK, CT. Data analysis: JH, DH, WW. Data interpretation: JH, CT, GRB, DH, ES. Drafting manuscript: JH, GRB, ES. Revising manuscript content: JH, VC, CT, CK, WW, GRB, DH, ES. Approving final version of manuscript: JH, VC, CT, CK, WW, GRB, DH, ES. JH, DH, ES take responsibility for the integrity of the data analysis.

