## Supplemental Documents for "Comprehensive Autoantibody Profiles in Systemic Sclerosis: Clinical Cluster Analysis"

***Supplemental Material***

| **Supp. Table 1.** Demographic, clinical, and serologic characteristics of 176 SSc patients with measured Anti-vascular receptor antibody, compared to full cohort | | | |
| --- | --- | --- | --- |
|  | | **AT_1_R and ET_A_R measured subgroup** | **All SSc patients** |
|  | | n = 176 | n = 372 |
| **Sex** | |  |  |
|  | No. (%) female | 144 (81.8) | 308 (82.9) |
|  | No. (%) male | 32 (18.2) | 64 (17.2) |
|  | Female-to-male ratio | 5:1 | 5:1 |
| **Age, mean ± SD – years** | |  |  |
|  | At onset RP | 44.76 ± 13.59 | 45.79 ± 16.85 |
|  | At diagnosis | 48.04 ± 15.12 | 47.99 ± 14.42 |
| **Disease duration, median (IQR) – years** | | 7.10 (10.80) | 7.45 (9.15) |
| **Disease classification, no. (%)** | |  |  |
|  | Diffuse (dcSSc) | 44 (25.0) | 104 (28.0) |
|  | Limited (lcSSc) | 120 (68.2) | 238 (64.0) |
|  | sine | 12 (6.8) | 30 (8.0) |
| **Antinuclear antibody (ANA) positive, no. (%)** | | 342 (95.3) | 342 (95.3) |
| **SSc manifestations, no. (%)** | |  |  |
|  | Raynaud symptoms | 158 (89.8) | 322 (86.6) |
|  | ILD | 74 (42.0) | 130 (34.9) |
|  | PAH | 24 (13.6) | 46 (12.4) |
|  | DU | 76 (43.2) | 133 (35.8) |
|  | Calcinosis | 25 (14.2) | 29 (8.0) |
|  | Cardiac involvement | 9 (5.1) | 8 (2.0) |
|  | Arthritis | 29 (16.5) | 33 (8.9) |
|  | SRC | 7 (4.0) | 10 (2.7) |
|  | Myositis | 10 (5.7) | 13 (3.0) |
|  | Terminal Organ failure | 5 (2.8) | 9 (2.4) |
|  | Malignancy | 14 (8.0) | 29 (7.8) |
| **Laboratory values,** median (IQR) or mean ± SD | |  |  |
|  | NT-proBNP – ng/L * | 126.00 (214.00) | 128.00 (216.00) |
|  | CRP – mg/dl * | 1.40 (4.20) | 0.75 (2.60) |
|  | Hb – mg/dl | 13.23 ± 1.58 | 11.30 ± 4.79 |
|  | Neutrophil granulocytes | 5.33 ± 2.63 | 5.38 ± 2.65 |
| **Cardiopulmonary parameters, mean ± SD** | |  |  |
|  | FVC – %pred | 89.65 ± 20.76 | 89.97 ± 20.29 |
|  | FEV1 - %pred | 86.62 ± 21.68 | 82.80 ± 28.65 |
|  | DLCO - %pred | 54.65 ± 17.58 | 52.91 ± 22.46 |
|  | LVEF - % | 61.8. ± 8.36 | 56.80 ± 16.92 |
| Disease duration refers to the time since first non-Raynaud symptom.  * median (IQR)  Abbreviations: CRP, C-reactive protein; dcSSc, diffuse cutaneous SSc; DLCO, diffusing capacity for carbon monoxide; FEV1, forced expiratory volume per second; FVC, forced vital capacity; Hb, hemoglobin; ILD, interstitial lung disease; IQR, interquartile range; L, liter; lcSSc, limited cutaneous SSc; LVEF, left ventricular ejection fraction; n, number; NT-proBNP, N-terminal-pro-brain natriuretic peptide; PAH, pulmonary arterial hypertension; RP, Raynaud’s phenomenon; SRC, scleroderma renal crisis; SSc, Systemic Sclerosis; %, percent; %pred, percent predicted | | | |

**Supp. Figure 1:**  Correlation Heatmap

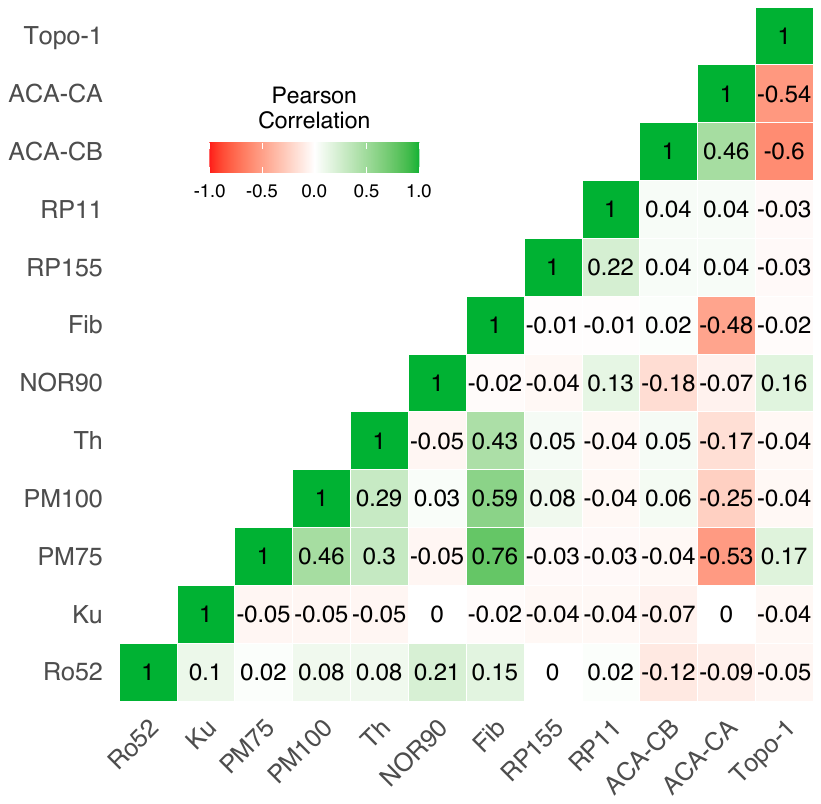

**Supp. Figure 1:**  Correlation Heatmap of SSc autoantibodies mesured by line blot.

**Supp. Figure 2:** Anti-vascular receptor antibodies

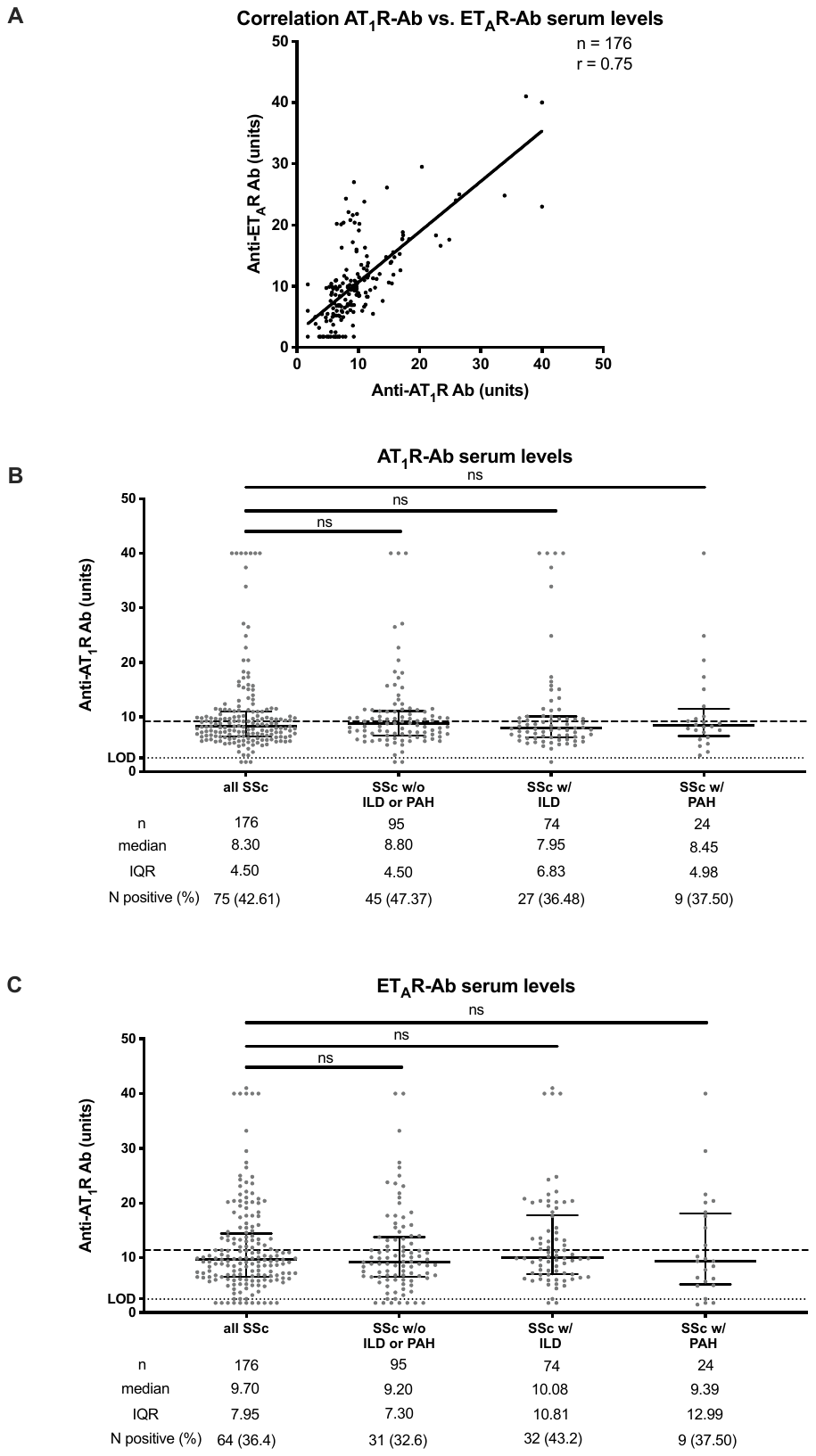

**Supp. Figure 2:** Anti-vascular receptor antibodies: Anti-AT_1_R and anti-ET_A_R autoantibody serum levels were evaluated in 176 SSc patients. Data are expressed as single values with median ± IQR. For significance tests Mann-Whitney-U test was used. The dashed line indicates the cut-off value. The dotted line indicates the lower detection limit of the ELISA. Values below the limit of detection (LOD) were shown as LOD/√2.
